# Brain Region-Centered MultiModal Hypergraph Fusion for MCI Conversion Prediction

**DOI:** 10.64898/2025.12.06.25341767

**Authors:** Zengjie Dong, Hong Liu, Xinting Ge, Hongkuan Zhang, Zhengxu Li, Yafan Chen, Wenhao Li

## Abstract

Alzheimer’s disease (AD) is a progressive neurodegenerative disorder, with mild cognitive impairment (MCI) as its prodromal stage. Accurate MCI conversion prediction is critical for early intervention and resource allocation. Recently, deep learning–based multi-modal neuroimaging fusion has become a hot research topic in AI-assisted AD diagnosis. Existing multimodal fusion approaches are limited by modality heterogeneity, ability to model inter-regional interactions, and insufficient interpretability. To address these challenges, BRC-MMHF, a Brain Region-Centered MultiModal Hypergraph Fusion framework, is proposed. In this framework, parameter-free channel exchange and ROI-level feature extraction mechanisms are employed to reduce modality heterogeneity and extract structurally consistent features from MRI and PET. A multimodal hypergraph models high-order interregional cross-modal relationships, while a lesion-aware module highlights disease-relevant regions to enhance interpretability. Structured clinical data are incorporated through a lightweight tabular encoder to improve adaptability and diagnostic robustness. Experiments on the ADNI dataset show that BRC-MMHF achieves 80.71% accuracy and 89.7% AUC in MCI conversion prediction, outperforming a range of state-of-the-art methods based on MRI and PET imaging, while providing high interpretability.

## I. Introduction

**A** LZHEIMER’S disease (AD) is a neurodegenerative disorder with complex pathophysiology, characterized by memory loss and irreversible cognitive decline [1]. By 2050, the number of individuals with AD is projected to exceed 100 million [2], posing a substantial burden on the families of patients and healthcare systems. Mild cognitive impairment (MCI) is the prodromal stage of AD, in which patients retain daily living abilities but already show significant cognitive decline, and early intervention can effectively slow disease progression [3]. However, not all MCI cases progress to AD. MCI can be classified as stable (sMCI) or progressive (pMCI) according to longitudinal follow-up, typically during a follow-up period of at least three years [4]. Prediction of such conversion types could support identification of highrisk individuals as well as provide guidance in personalized interventions and resource allocation.

Accurate MCI conversion prediction requires multimodal analysis. Beyond cognitive scales and tests, imaging is crucial: structural MRI (sMRI) detects brain atrophy and lesions and is strongly linked to AD pathology [5], while positron emission tomography (PET) measures metabolic decline or *β*-amyloid deposition, providing a key tool for early detection of AD-related changes [6]. With advances in deep learning, researchers have used convolutional neural networks (CNNs) to extract features from MRI or PET images, capturing subtle brain changes for automated diagnosis [7]–[9]. However, single image modality captures only one aspect of brain structure or function, limiting the ability to fully characterize AD pathology and achieve accurate and robust MCI conversion prediction.

Combining MRI and PET to capture comprehensive brain features has become a popular strategy in recent studies, which can address the limitations of single image modality [10], [11]. However, multimodal fusion remains challenging, as most existing studies employ simple strategies such as feature concatenation [12], [13], which fail to achieve effective cross-modal feature alignment, resulting in modality heterogeneity and reduced fusion performance. Recent studies have employed transformer and cross-attention mechanisms [14], [15] to mitigate modality heterogeneity. However, these mechanisms often require high computational cost and lack regional modeling, and hence offer limited interpretability. Another line of work uses graph neural networks (GNNs) to model brain regions at the region of interest (ROI) level [16]–[18], where features from each ROI are often aggregated into a single representative statistic. This approach is computationally efficient and inherently interpretable, but loses spatial information, and hence limiting its feature representation and prediction accuracy.

To address these challenges of multimodal image fusion and interpretability in MCI conversion prediction, we propose BRC-MMHF, a Brain Region-Centered MultiModal Hyper-graph Fusion framework. It aligns and integrates MRI and PET data for comprehensive brain analysis and leverages hyper-graph modeling to capture high-order inter-regional relationships and enhance interpretability. Specifically, BRC-MMHF uses a parameter-free channel exchange mechanism in the imaging feature extraction stage, which enhances multimodal feature representation and alleviates modality heterogeneity. An ROI-based strategy extracts structurally aligned brain region features to construct a multimodal brain region hypergraph. Hyperedges explicitly model high-order relationships between regions, enhancing regional modeling capacity. A lesion-aware module identifies key brain regions, providing interpretability for the prediction results. In addition, to ensure better coherence with clinical practice and to enhance usability, BRC-MMHF integrates a structured clinical data analysis module into the fusion framework. This module employs a lightweight tabular encoder to efficiently model phenotypic features and integrate them with imaging features, further improving the framework’s adaptability to individual differences and its accuracy of diagnostic prediction.

Compared with existing approaches, particularly those based on transformer or GNNs architectures, BRC-MMHF provides a more comprehensive solution for multimodal image fusion. It adopts a progressive information flow, from channel exchange to brain region-level hypergraph fusion, to align and enhance MRI and PET features while modeling cross-modal brain region associations, thereby mitigating modality heterogeneity and improving regional modeling. In addition, BRC-MMHF leverages hypergraph modeling to capture high-order associations, overcoming the binary-edge, pairwise limitations of GNNs, while the hypergraph attention mechanism further strengthens interpretability. The framework is validated on the publicly available ADNI dataset using sMRI, FDG PET, and structured clinical data.

Overall, the main contributions of this work can be summarized as follows:

- A Brain Region–Centered MultiModal Fusion Framework (BRC-MMHF) is proposed, in which MRI and PET data are progressively integrated to mitigate modality heterogeneity, capture inter-regional relationships, and enhance disease-relevant modeling. Additional structured clinical data are processed to improve adaptability and diagnostic robustness.
- A multimodal hypergraph with attention is introduced to explicitly capture high-order inter-regional associations of brain regions, thus overcoming the pairwise limitations of conventional graph-based methods.
- A lesion-aware node scoring mechanism is designed to identify clinically relevant brain regions, thereby improving the interpretability of diagnostic predictions.
- Comprehensive experiments on the ADNI dataset are conducted, and the results demonstrate that BRC-MMHF outperforms recent state-of-the-art methods, while providing high interpretability and clinical applicability.

The remaining sections of this paper are organized as follows: Section II reviews related work. Section III describes the proposed framework. Section IV presents experiments on a public dataset. Section V discusses study limitations and future directions. Section VI concludes the paper.

## II. Related work

### A. Multimodal MCI Conversion Prediction

Traditional MCI conversion prediction approaches process clinical and imaging data through feature engineering or radiomics, combined with empirical thresholds and classical machine learning models. For example, some methods use MMSE, ADAS, and demographic information [19], while imaging-based studies apply voxel-based morphometry (VBM) to extract features such as gray matter volume and cortical thickness [20]–[23]. In recent years, with the rapid development of CNNs and other deep learning techniques, their application in early AD diagnosis has demonstrated significant advantages. Representative approaches use 3D CNN models to extract features from MRI or PET images, achieving better performance in MCI conversion prediction than traditional machine learning [7]–[9]. However, reliance on a single imaging modality limits the representation of disease information, reducing prediction accuracy and robustness.

Previous studies have shown that multimodal data fusion, especially multiple imaging modalities, provide complementary neuropathological information and enhance the identification and prediction of MCI progression [10], [24]. Recently, more studies have focused on multimodal imaging fusion to improve prediction performance and interpretability through modality complementarity. Huang et al. [11] used 3D-VGG to extract features from MRI and PET of the hippocampal region and concatenated them. Some studies have directly concatenated MRI and PET in the channel dimension for early fusion [12], [13], showing improved performance. However, these simple strategies ignore modality heterogeneity, namely imaging differences between modalities, which cause inconsistencies in feature distributions and semantic spaces, weakening fusion effectiveness. For example, MRI captures structural changes while PET highlights metabolic abnormalities, leading to heterogeneous feature distributions and semantic misalignment that hinder effective fusion when modalities are naively concatenated. To address modality heterogeneity, some studies have introduced cross-modal interaction at the feature layer [14], [25], [26], where independent encoders are used to extract features from each modality, and crossattention mechanisms are applied to achieve information complementarity. The transformer framework, as a representative approach in this category, has shown strong fusion capability and achieved state-of-the-art performance in MCI conversion prediction [15], [27], [28]. However, these methods rely on CNNs to extract whole-brain features and then feed them into transformer modules. Therefore, they generally lack brain regional modeling and include substantial irrelevant noise, which limits their interpretability and increases computational cost.

### B. Graph-based Feature Fusion

Recent studies have used GNNs for multimodal feature alignment and fusion in MCI conversion prediction [16]– [18], [29]. Graphs represent brain regions as nodes and interregional relationships as edges, with associations modeled via edge weights or learnable adjacency matrices for ROI-level fusion. Leveraging the advantages of graph structures, this approach highlights key regions, reduces noise, and improves interpretability. However, these GNN-based methods are limited to pairwise graphs and cannot represent multiregion lesion patterns, such as simultaneous abnormalities in the bilateral hippocampus and parahippocampal gyrus, which require higher-order modeling. Meanwhile, hypergraphs have strong higher-order relationship modeling capabilities [30]. With capability of representing higher-order interactions across brain regions and modalities, hypergraphs can better characterize complex pathological patterns. Building on this advantage, this study replaces traditional graphs with a brain region-level multimodal hypergraph to capture higher-order associations, thereby enhancing the representational capacity and predictive performance of the diagnostic model.

### C. Diagnostic Interpretability

Beyond predictive accuracy, interpretability is another key consideration in MCI conversion prediction, as it provides evidence-based clinical insight into disease mechanisms. Given the complex pathology of Alzheimer’s disease, interpretability is particularly important for revealing the mechanisms underlying MCI progression [17], [31]. Medical interpretability techniques, such as class activation mapping (CAM) and Grad-CAM, visualize activation patterns to explain features relevant to model outputs [32]. However, most methods rely on CNN-based heatmaps of 3D whole-brain images, lacking brain regional modeling and importance analysis [26], [33]. High-performance models often sacrifice interpretability, whereas interpretability-focused methods may underperform [34]. One of the main challenges in MCI conversion prediction is to design a framework that effectively balances predictive accuracy and interpretability.

### D. Comparison with Existing Approaches

Existing multimodal image fusion approaches for MCI conversion prediction are generally classified into two categories: Global feature-based methods, which integrate MRI and PET features at the whole-brain level, and Region-based methods, which model inter-regional associations through graph-based frameworks. While both methods mitigate modality heterogeneity and enhance regional modeling, they suffer from inherent limitations, such as inadequate cross-modality alignment and an inability to capture higher-order inter-regional relationships.

Compared with existing multimodal image fusion strategies and approaches, BRC-MMHF introduces a region-centered hypergraph framework that captures higher-order associations across multiple brain regions and different modalities. It employs a parameter-free channel exchange to mitigate modality heterogeneity and utilizes hypergraphs to model associations among brain regions. The hypergraph structure enables representation of multi-region lesion patterns beyond pairwise connections, while hypergraph attention highlights disease-relevant regions and strengthens interpretability. In summary, BRC-MMHF provides an integrated fusion framework that improves modality alignment and regional interpretability, offering a more reliable solution for MCI conversion prediction.

## III. Method

This study proposes BRC-MMHF, a region-centric multimodal hypergraph fusion framework that aligns and integrates MRI and PET data for comprehensive brain structure analysis in downstream tasks. The overall workflow of BRC-MMHF is shown in Fig. 1.

**Fig. 1:**
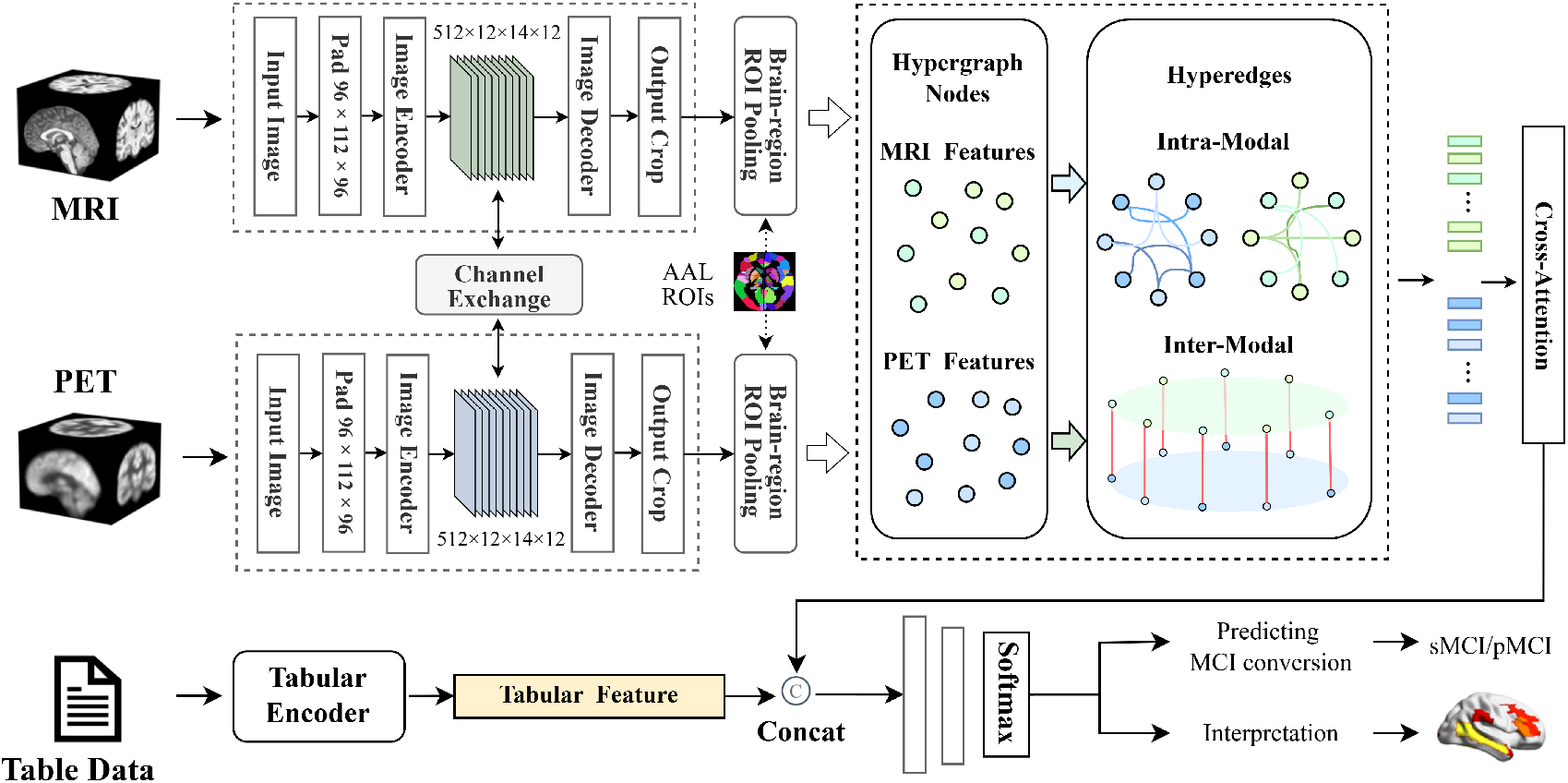
The overall framework includes the imaging and non-imaging branches, along with the MCI conversion prediction and interpretability analysis tasks. The BRC-MMHF framework proposed in this paper is the core of the imaging branch, including dual-modal imaging feature extraction and hypergraph-based brain region-level multimodal fusion.

### A. Problem Formulation

Given a multimodal dataset 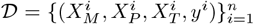, where 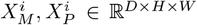 denote the preprocessed MRI and PET images of subject *i*, and 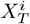 represents the structured clinical data, the objective is to learn a mapping function:

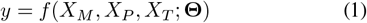

where **Θ** denotes the trainable parameter set. The function *f* performs multimodal feature extraction and fusion to predict *y*^*i*^ ∈ {0, 1}, indicating MCI-to-AD conversion within 36 months (sMCI vs. pMCI).

In the multimodal feature extraction stage, the structured data *X*_*T*_ is fed into a lightweight tabular encoder to obtain the encoded representation: **z**_*T*_ = *TabE*(*X*_*T*_). MRI and PET data are processed by a 3D feature extractor to extract brain-region-level feature representations:

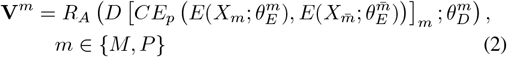

where *E*(·) and *D*(·) denote the encoder and decoder in the feature extraction stage. *X*_*m*_ and 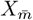 are the dual-modality image inputs, and 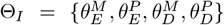 is the set of parameters of the feature extractor. The operator *CE*_*p*_, parameterized by a channel-exchange ratio *p*, performs channel exchange at the encoder’s deepest layer. This parameter-free operation leverages the scaling coefficients from Batch Normalization (BN) and introduces no additional learnable parameters, thus requiring no extra training. The function *R*_*A*_(·) is an ROI feature extractor based on the brain atlas *A* ∈ {1, …, *N*} ^*D*×*H*×*W*^, producing brain-region-level features **V**^*m*^ ∈ ℝ^*N* ×*C*^ for both modalities, where *N* is the number of brain regions and *C* is the number of feature channels.

A hypergraph is defined as 𝒢 = (𝒱, ℰ), where the node set 𝒱 = *v*_1_, *v*_2_, …, *v*_*n*_ represents entities or concepts. The hyperedge set ℰ = *e*_1_, *e*_2_, …, *e*_*m*_ connects multiple entity nodes to represent complex high-order relationships. Each hyperedge *e*_*j*_ ⊆ 𝒱 is associated with a set of nodes. The relationship matrix *H* ∈ℝ^*n*×*m*^ describes the high-order relations between *m* hyperedges and *n* nodes. In the context of this study, brain region features and inter-regional relationships are modeled as follows: Hypergraph nodes represent brain region–level features, with initial node features denoted as **X**^(0)^ = [**V**^*M*^ **V**^*P*^] ∈ ℝ ^(2*N*)×*C*^. Hyperedges connect multiple functionally related or structurally adjacent brain region nodes to model their higher-order relationships. The node feature update process can be expressed as:

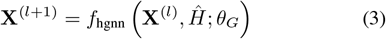

where *f*_hgnn_ denotes the function that updates the node features using the dynamic hypergraph attention mechanism. *Ĥ* = [*Ĥ*_*M*_; *Ĥ*_*P*_] denotes the dynamic hypergraph incidence matrix updated via the hypergraph attention mechanism, and *θ*_*G*_ is the set of hypergraph learning parameters.

The updated multimodal brain region features 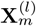 pass through a cross-attention module to aggregate dual-imaging feature representations **z**_*m*_, then fuse with structured data features before entering the classifier for prediction:

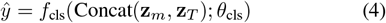

where *Concat*(·) denotes the feature concatenation operation. *f*_cls_ is a classifier composed of multiple fully connected layers, performing the MCI conversion prediction task, and *θ*_cls_ represents the classifier parameters. Binary cross-entropy loss is used during training:

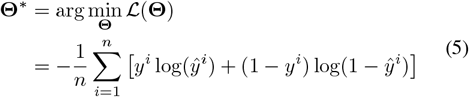

where **Θ** = {*θ*_*I*_, *θ*_*G*_, *θ*_cls_} denotes the set of trainable parameters, *n* is the number of samples, *y*^*i*^ is the ground truth label of sample *i*, and *ŷ*^*i*^ is the predicted probability.

### B. Image Branch

#### 1) Multimodal Imaging Feature Extraction

BRC-MMHF employs an improved 3D U-Net to extract brain region-level features from MRI and PET. The encoder-decoder architecture with skip connections captures multi-scale contextual information while preserving spatial details, enabling high-resolution feature reconstruction [35]. In the decoder, an ROI-based extraction strategy guided by a brain atlas outputs brain region-level features and forms the foundation for subsequent multimodal brain region hypergraph fusion. Unlike directly using 3D U-Net for classification [14], BRC-MMHF introduces segmentation supervision to guide the network in learning anatomically consistent regional features, improving sensitivity to brain region differences and enhancing the discriminative power and stability of downstream models. This approach has been validated in multiple medical imaging studies [36] and is particularly suitable for AD identification tasks with highly localized lesion features.

As shown in Fig. 2, the network comprises multiple encoder layers and symmetric decoder layers. Each encoder layer performs two 3D convolutions followed by max pooling for spatial downsampling, doubling the number of channels at each level. The decoder upsamples the deep features using 3D transposed convolutions, restores spatial resolution, and concatenates them with the corresponding skip-connected features 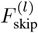 from the encoder along the channel dimension, followed by two 3D convolutions for feature fusion.

**Fig. 2:**
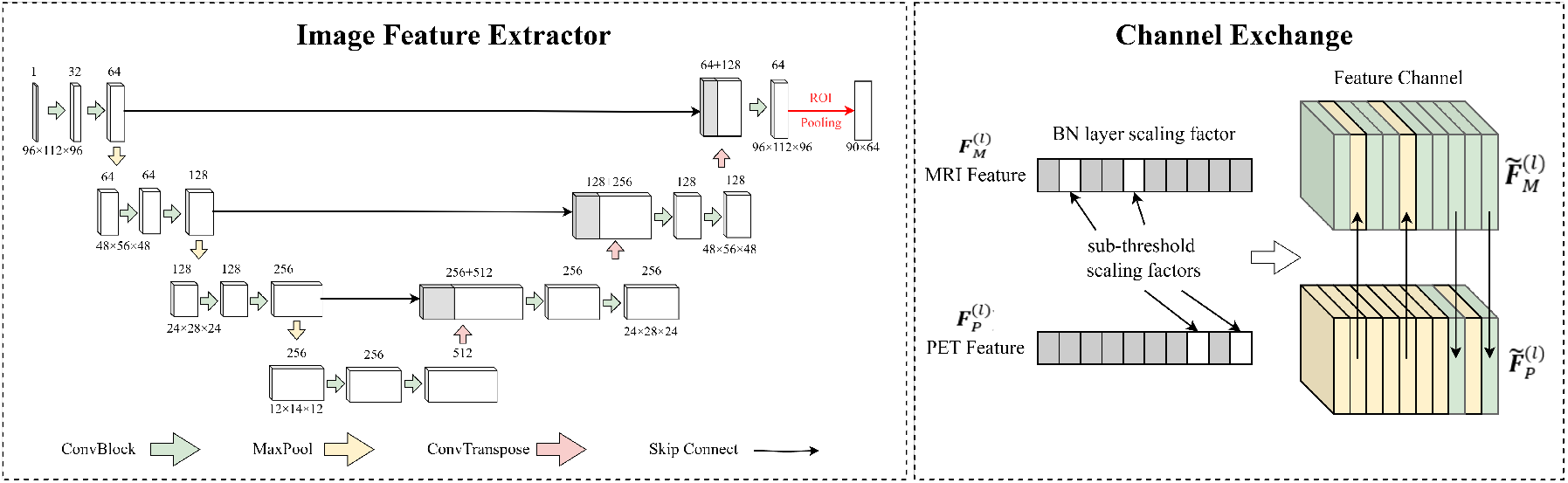
Image encoder: Left panel shows the 3D U-Net feature extractor with a shared bottleneck and ROI pooling for deriving brain region-level features; right panel illustrates channel exchange between MRI and PET features at the shared layer.

#### 2) Cross-Modal Channel Exchange

The deepest layer of the encoder is the bottleneck, which facilitates cross-modal feature interaction to reduce modality differences. BRC-MMHF introduces a channel exchange mechanism in the bottleneck layer. In the bottleneck, the features of MRI and PET are denoted as 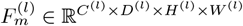, where *m* ∈ {*M, P*} indicates the modality. At the BN layer, the absolute value of the scaling factor *γ*_*m,c*_ measures the contribution of channel *c* to the current modality. As shown in Fig. 2, channels are ranked by |*γ*_*m,c*_|, exchange weak channels one-to-one across modalities, and keep strong channels unchanged. The exchanged features are formulated as:

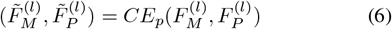

where *p* is the channel exchange ratio controlling the proportion of channels exchanged across modalities. The exchanged channel set is adaptively updated during training. If a previously weak channel becomes stronger, its |*γ*| increases and the channel will be removed from the exchange set in the next iteration. This process uses the existing trainable parameters *γ* and introduces no extra parameters, which allows for seamless integration into deep feature fusion.

Shared bottleneck parameters are applied to produce semantically consistent two-modal features for semantic alignment purposes of the mechanism. Considering the information bottleneck principle [37], the bottleneck layer of the 3D U-Net is chosen for parameter sharing and channel exchange, as it is the transition point from modality-specific to shared semantic representations. Early layers focus on modality-specific lowlevel features. Exchanging them too early may introduce cross-modal noise. In contrast, the bottleneck stage provides compact high-level semantic representations, where modality discrepancies are mainly distributional rather than structural, making it the best point for selective feature alignment. In later decoding stages, the ability to correct modality gaps becomes limited, since features have already been aggregated and the decoder primarily restores spatial resolution rather than re-aligning distributions. In summary, the bottleneck layer combines discriminative capability with abstraction, fusing high-level cross-modal information while preserving modality-specific characteristics. Previous work [38] has also shown that sharing parameters at this position achieves optimal fusion and helps the decoder recover spatial structures more accurately.

#### 3) Hypergraph Construction

After channel exchange, ROI pooling is performed to ensure that features from different modalities correspond to the same anatomical regions. This anatomical alignment avoids spatial mismatch, guarantees semantic consistency across nodes for hypergraph construction, and reduces the search space of cross-modal interactions, thereby making region-level modeling more stable and interpretable.

At the decoder output, the feature map *F*_out_ is cropped to the original spatial size: 91 × 109 × 91 with a voxel size of 2 mm^3^, corresponding to the standard 2 mm^3^ MNI space. The ROI atlas is denoted as *A* ∈ ({1, …, *N*})^*D*×*H*×*W*^, where *N* is the number of brain regions. The Automated Anatomical

Labeling (AAL) atlas [39] is applied, which contains regions labeled 0-116. Label 0 indicates undefined background, and 26 small cerebellar regions are excluded from feature extraction, resulting in *N* = 90. Based on this atlas, ROI-level features are computed for each modality as follows: let Ω_*r*_ denote the set of voxel coordinates in brain region *r*, Ω_*r*_ = {(*x, y, z*) | *A*(*x, y, z*) = *r}*. For each channel *c* ∈ {1, …, *C}*, the average response in region *r* is computed as:

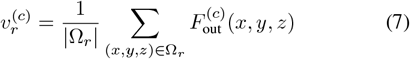

where 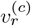 is the mean value of channel *c* in region *r*, |Ω_*r*_ | is the voxel count in *r*. The features from all channels in region *r* constitute a vector: 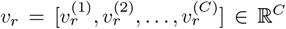. For modality *m*, the ROI-level feature 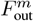 is extracted according to atlas *A*:

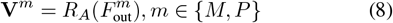

Further, the brain region–level feature for each modality is **V**^*m*^ = {*v*_*m,r*_ | *m* ∈ {*M, P*}, *r* ∈ {1, 2, …, *N*}} ∈ ℝ ^*N* ×*C*^ which is used as the node representation in the hypergraph.

The hypergraph is defined as 𝒢 = {𝒱, ℰ}. The node set is 𝒱= {**V**^*m*^ | *m* ∈ {*M, P*}}, representing brain region–level features from both MRI and PET modalities. The hyperedge set ℰ contains two types: ℰ _intra_ for intra-modal connections and ℰ _inter_ for inter-modal connections. ℰ _intra_ connect different brain regions within the same modality to capture high-order intra-modal relationships, while ℰ _inter_ link the MRI node *v*_*M,r*_ and PET node *v*_*P,r*_ of brain region *r* to capture disease-related cross-modal relationships. When constructing, each hyperedge links a node to its *top* − *k* nearest neighbors based on cosine similarity in the feature space.

The node–hyperedge incidence matrix is denoted as *H* ∈ ℝ ^| 𝒱 |×|ℰ |^. For intra-modal connections, *H*_*M*_ and *H*_*P*_ denote the brain-region relation matrices for MRI and PET, respectively. For inter-modal connections, *H*_inter_ encodes cross-modal relations weighted by feature importance. We concatenate these matrices as *H* = [*H*_*M*_ ∥*H*_*P*_∥ *H*_inter_]. Unlike traditional graphs that connect only two nodes, the hypergraph allows a hyperedge to simultaneously connect multiple nodes, enabling a richer representation of high-order associations [30], [40].

#### 3) Hypergraph Fusion

Although conventional GNNs are effective for capturing pairwise dependencies, they fall short in explicitly modeling multi-region interactions as unified entities [14], [16]. Hypergraph learning overcomes this limitation by using hyperedges to directly connect multiple ROIs, thereby representing high-order relationships. This enables the model to capture co-occurring lesion patterns.

The initial node features are denoted as **X**^(0)^. Node features and hypergraph structure are input to a hypergraph convolution layer (HGNNConv^+^) [41], where messages passing between nodes and hyperedges are:

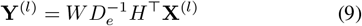

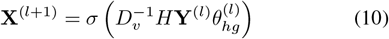

where *D*_*v*_ is the vertex degree matrix, *D*_*e*_ is the hyperedge degree matrix, and *W* is a learnable weight matrix. **Y**^(*l*)^ denotes aggregated features from nodes to hyperedges, while **X**^(*l*+1)^ represents updated node features after message passing from hyperedges. Node features for each modality are updated using *f*_hgnn_(**X**^(*l*)^, *H*_*M*_) and *f*_hgnn_(**X**^(*l*)^, *H*_*P*_) to capture high-order intra-modal relationships.

Since hypergraph convolution inherently incorporates an internal attention mechanism (Eq. (9)), it weights incoming and outgoing information at each node by learned importance. However, these weights are fixed after training and cannot adapt dynamically. Therefore, we introduce a hypergraph attention mechanism [42] to learn a dynamic incidence matrix Ĥ = *f*_hatt_(**X**^(*l*)^, (*H*)^⊤^), where the element *ĥ*_*v,e*_ is defined as: 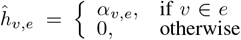. Here, *α*_*v,e*_ is a learnable dynamic attention coefficient, measuring the importance of node *v* to hyperedge *e*, calculated as:

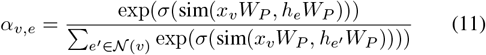

where *x*_*v*_ is the brain region feature vector of node *v*, and *h*_*e*_ is the aggregated feature of hyperedge *e. W*_*P*_ is the trainable projection matrix mapping node and hyperedge features into the same semantic space. 𝒩 (*v*) denotes hyperedges incident to node *v*. sim(·) is the similarity function defined in [42]. The learned dynamic incidence matrix *Ĥ* replaces *H* in Eqs. (9) and (10) to build adaptive brain-region relationship graphs.

For inter-modal hyperedges, to enhance the interaction between MRI and PET features, cross-modal attention is applied on the inter-modal incidence matrix *H*_inter_ to fuse features of the two imaging modalities. Since *H*_inter_ is constructed per brain region, cross-attention is computed locally for each region, reducing complexity while still enabling effective fusion of features from both image modalities.

This hypergraph-based design not only strengthens intra- and inter-modal interactions, but also forms a key component of the overall BRC-MMHF pipeline. BRC-MMHF first reduces heterogeneity at a semantically rich stage (bottleneck), then grounds features in a common spatial framework (ROI alignment), and finally models them in a structure capable of high-order interactions (hypergraph), achieving a balanced trade-off among modality alignment, structural consistency, and relationship expressiveness. This progression also facilitates subject-specific interpretability via the Lesion-Aware Node Scoring (LANS, Section III-E) module.

### C. Tabular Branch

In the clinical assessment of Alzheimer’s disease, structured data such as clinical diagnoses, cognitive tests, and biomarkers contain rich information that is highly valuable for early AD diagnosis and MCI conversion prediction. However, their diversity and frequent missing values nature hinder effective feature extraction. Traditional approaches, such as one-hot encoding and normalization, fail to capture cross-column interactions, and small-sample settings are prone to severe overfitting, ultimately impacting downstream task performance.

To address these challenges, inspired by the TabPFN framework [43], a tabular encoder is designed, as illustrated in

Fig. 3. The encoder comprises two parallel branches: **Feature Attention**, which captures feature dependencies across columns, and **Sample Attention**, which captures sample dependencies across rows. For a patient’s structured clinical data 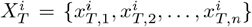, a learnable embedding vector is first generated for each column 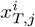. For categorical data, categories are mapped to integer indices and then converted into vectors via a lookup embedding matrix *E*_cat_. Continuous data are first *z*-score normalized, followed by a learnable linear transformation matrix *W*_cont_ that maps the standardized scalar to an embedding vector. For missing values, a dedicated embedding vector *e*_*m*_ with the same dimensionality is generated.

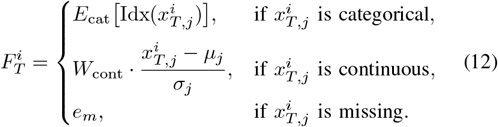

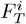 is fed into a lightweight Transformer encoder to obtain the feature representation **z**_*T*_. Subsequently, **z**_*T*_ can be fused with imaging-modality features or independently passed to the classification layer to predict MCI conversion.

**Fig. 3:**
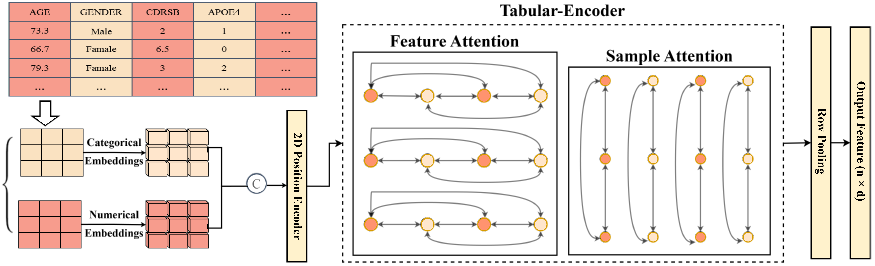
Tabular Encoder: Applies row and column attention mechanisms to process structured clinical data and generate encoded features.

**Fig. 4:**
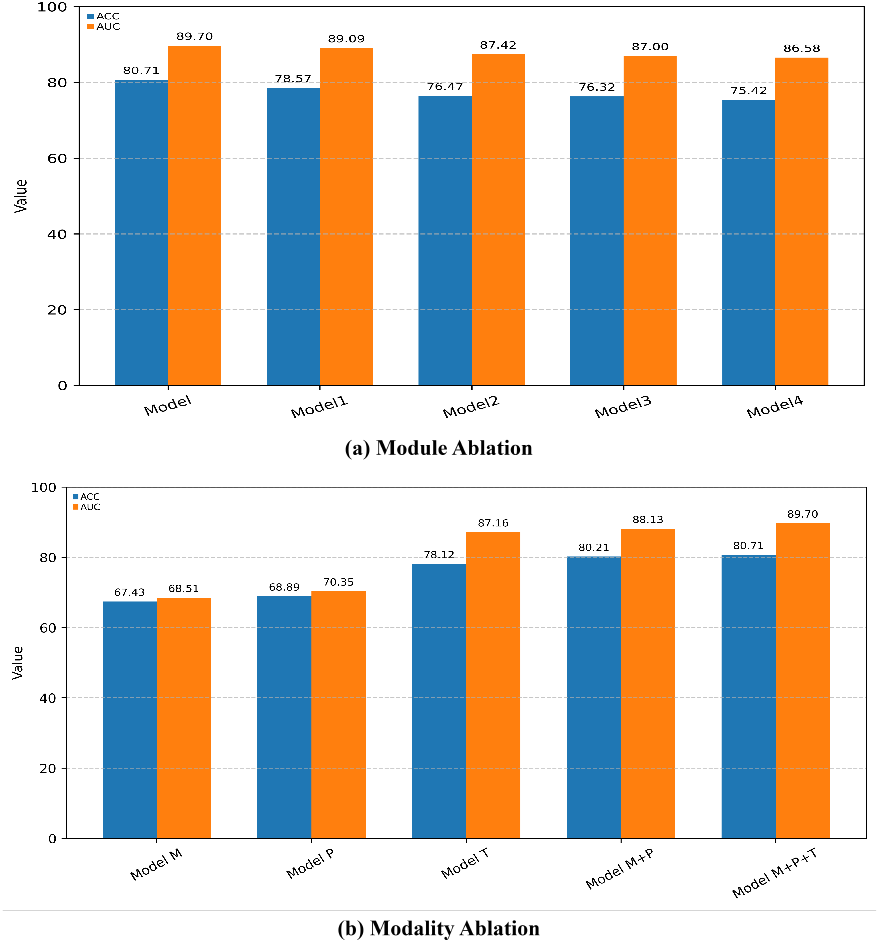
Bar chart of ablation experiment results.

### D. Output

The output features **z**_*T*_ from the tabular branch are concatenated with the output features **z**_*m*_ from the imaging branches for final fusion, representing the patient’s overall condition: 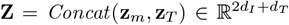, where *d*_*I*_ and *d*_*T*_ are the dimensions of imaging and tabular features. The classifier is a multi-layer fully connected network, followed by a softmax activation to produce the classification result:

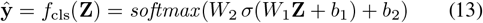

where *W*_1_, *W*_2_ and *b*_1_, *b*_2_ are learnable parameters of the fully connected layers.

### E. Lesion-Aware Node Scoring

After computing the hypergraph attention *Ĥ*, the attention score *α*_*v,e*_ between node *v* and hyperedge *e* is obtained. Then, a Lesion-Aware Node Scoring (LANS) module is placed to evaluate the importance of each brain region by averaging the attention scores across its connected hyperedges:

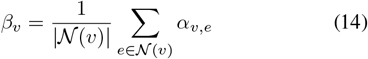

where 𝒩 (*v*) denotes all hyperedges incident to node *v*. Furthermore, the top-*k* brain regions for each patient is identified to provide interpretable clinical insights. The BRC-MMHF framework extracts and analyzes brain region-level features, constructs graphs based on AAL-defined regions, and fuses multimodal information. During inference, the attention weights directly highlight critical regions, enhancing clinicians’ trust in AI-assisted diagnosis.

## IV. Experiments

### A. Experimental Data

#### 1) Dataset

The data used in this study were obtained from the Alzheimer’s Disease Neuroimaging Initiative (ADNI), a database widely used for international Alzheimer’s disease research (https://ida.loni.usc.edu). The data selection followed the majority of previous studies on MCI conversion prediction [15], [33]. The time window was set to 36 months. Participants whose condition progressed to dementia in this period were labeled pMCI; others were labeled sMCI. As Alzheimer’s disease is irreversible, cases with diagnostic reversion were excluded. We enrolled 479 participants, including 321 sMCI and 158 pMCI cases. Their demographic information is shown in Table II.

**TABLE 1:**
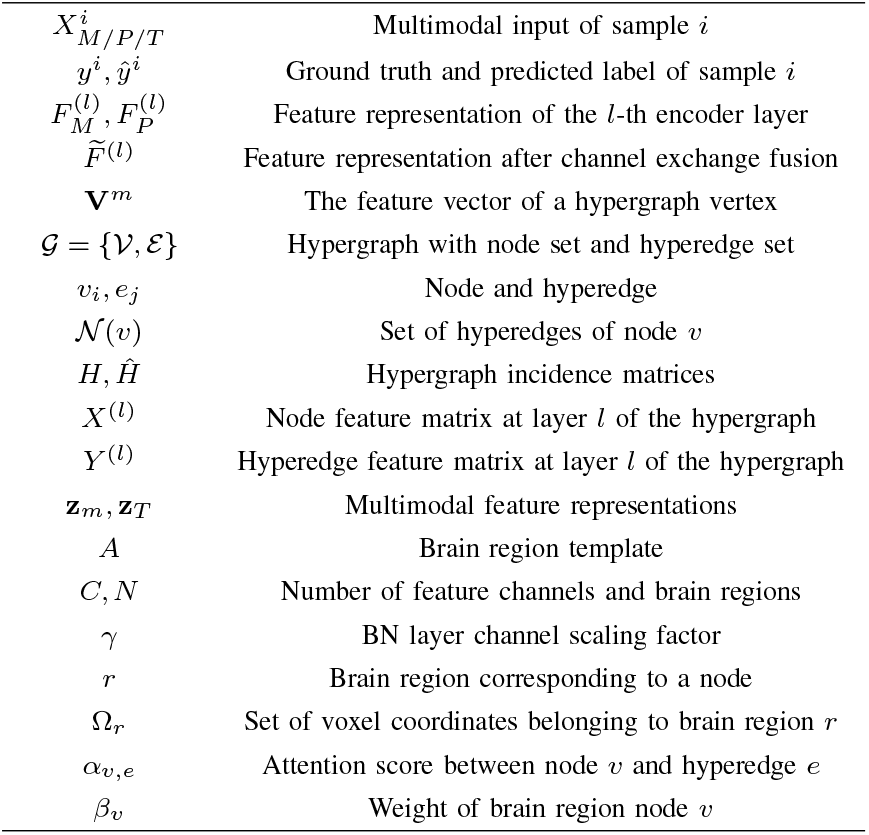
Notations and Descriptions.

**TABLE 2:** Demographic information of participants.

| Category | sMCI | pMCI |
| --- | --- | --- |
| Number | 321 | 158 |
| Age | 72.49 ± 7.53 | 73.62 ± 6.94 |
| Gender | 133/188 | 64/94 |
| Education | 16.01 ± 2.79 | 15.97 ± 2.63 |
| MMSE | 27.98 ± 1.73 | 27.13 ± 1.75 |
| CDR-SB | 1.3 ± 0.73 | 1.95 ± 0.98 |
| FAQ | 1.96 ± 3 | 5.22 ± 4.32 |
| APOE (0/1/2) | 187/103/28 | 49/80/29 |

MMSE, Mini-Mental State Examination; CDR-SB, Clinical Dementia Rating—Sum of Boxes; FAQ, Functional Activities Questionnaire; APOE, Apolipoprotein E (with allele types 0, 1, and 2 representing different genetic variations).

#### 2) Data Preprocessing

Image preprocessing followed the ADNI guidelines (https://ida.loni.usc.edu/). Specifically: (1) MPRAGE multiplanar reconstruction; (2) 3D GradWarp correction; (3) B1 field inhomogeneity correction; (4) N3 intensity normalization. MRI images were then AC-PC aligned, skull-stripped, and registered to the MNI152 space with 2 mm voxels. For PET images, after AC-PC alignment, MRI-derived (13) deformation fields were applied for precise registration.

For structured clinical data, we used the ADNI database from the ARC Builder on the USC IDA platform to obtain clinical assessments and biomarker results, aligned with corresponding imaging dates. To reflect AD clinical practice, we focused on in total of 133 non-imaging variables from interviews, physical exams, and cognitive tests.

### B. Experimental Setup and Evaluation Metrics

This experiment employed 5-fold cross-validation, dividing the dataset into five non-overlapping subsets. In each iteration, one subset was used for testing and the remaining four for training. This strategy prevents data leakage, ensures result consistency and reliability, and reports the final performance as the mean across all folds.

The experimental framework was implemented in PyTorch (version 2.4.1) and executed on a workstation equipped with an NVIDIA GeForce RTX 3090 GPU with 24 GB of memory. Training used the Adam optimizer (1 × 10^−6^ learning rate, batch size 4, weight decay 1 × 10^−4^) with cosine annealing for 100 epochs and cross-entropy loss. 3D images were zero-mean/unit-variance normalized, with random Gaussian noise augmentation during training and normalization only for validation and testing. In the experiments, the channel exchange ratio *p* was set to 0.2, and two hyperedge scales of *k* = 6 and *k* = 18 were used to capture both local and global structures. The image feature dimension *d*_*I*_ was set to 512, the tabular feature dimension *d*_*T*_ was set to 192, and the final feature vector **Z** for classification had a dimension of 1216. The experimental settings are detailed in Table III.

**TABLE 3:**
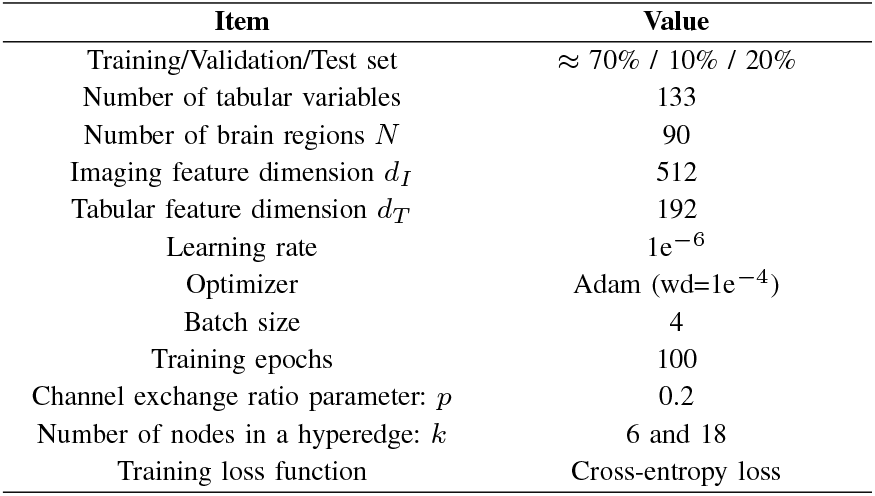
Experimental parameter settings.

| Item | Value |
| --- | --- |
| Training/Validation/Test set | $\approx 70\% / 10\% / 20\%$ |
| Number of tabular variables | 133 |
| Number of brain regions $N$ | 90 |
| Imaging feature dimension $d_I$ | 512 |
| Tabular feature dimension $d_T$ | 192 |
| Learning rate | $1e^{-6}$ |
| Optimizer | Adam (wd=$1e^{-4}$) |
| Batch size | 4 |
| Training epochs | 100 |
| Channel exchange ratio parameter: $p$ | 0.2 |
| Number of nodes in a hyperedge: $k$ | 6 and 18 |
| Training loss function | Cross-entropy loss |

Multiple metrics are applied for evaluation of the model’s performance, including accuracy (ACC), sensitivity (SEN), specificity (SPE), and area under the curve (AUC), to comprehensively assess classification capability.

### C. Comparative Experiments

To validate the effectiveness of the proposed BRC-MMHF framework for MCI conversion prediction, we compared it with recent state-of-the-art methods on sMCI vs. pMCI classification, including convolutional, Transformer, and graph neural network approaches with unimodal and multimodal inputs on the ADNI dataset. Comparison methods are listed as follows:

- 3D-VGG: Silvia Basaia et al. (2019) proposed a method using deep neural networks to process MRI images for MCI conversion classification [7].
- ROI-3D CNN: Yechong Huang et al. (2019) proposed a ROI-based 3D Convolutional Neural Network for processing MRI and PET data [11].
- 3D DenseNet: Jie Zhang et al. (2021) proposed a 3D DenseNet with connection-wise attention to process MRI imaging data [8].
- CF-3D CNN: Zhaokai Kong et al. (2022) proposed a early channel fusion multimodal method based on 3D CNN, utilizing MRI and PET data [13].
- Aging-Transformer: Xingyu Gao et al. (2023) proposed a framework combining multi-scale attention and Transformer architectures using structural MRI data [44].
- DAUF: Zhehao Zhang et al. (2023) proposed a U-Net-based method with attention mechanisms for MCI conversion prediction using structural MRI data [9].
- MMCA: Jin Zhang et al. (2023) proposed a multimodal fusion method based on cross-attention mechanisms to integrate MRI, PET, and CSF structured data [14].2
- AFSNet: Jinwei Liu et al. (2024) proposed an attention-based method with lesion feature selection to process MRI data at multiple time points for MCI conversion prediction [33].
- SGCN: Houliang Zhou et al. (2025) proposed a graph convolutional network (GCN)-based method to fuse MRI and PET data [18].
- MF-Transformer: Yuanwang Zhang et al. (2025) proposed a Transformer-based framework to integrate MRI, PET, and structured clinical data [15].

As shown in Table IV, our method achieved 80.71%, 72.73%, 92.68%, and 89.7% in ACC, SEN, SPE, and AUC, respectively, representing improvements of 2.17% and 4.10% in ACC and AUC over recent state-of-the-art methods. The large discrepancy between SEN and SPE is mainly due to the imbalance between sMCI and pMCI samples. By calculating the balanced accuracy (BACC = (SEN + SPE)*/*2), which eliminates the impact of sample imbalance on accuracy, our method achieved a BACC of 82.71%, outperforming all other methods.

**TABLE 4:** Comparison with recent state-of-the-art methods (sMCI vs. pMCI on ADNI).

| Model | Data Used | ACC(%) ↑ | SEN(%) ↑ | SPE(%) ↑ | BACC(%) ↑ | AUC(%) ↑ |
| --- | --- | --- | --- | --- | --- | --- |
| 3D VGG (2019) | MRI | 69.24 | 62.75 | 79.30 | 71.03 | 75.43 |
| ROI-3D CNN (2019) | MRI+PET | 72.22 | 68.15 | 83.93 | 76.04 | 79.61 |
| 3D DenseNet (2021)* | MRI | 78.79 | 75.16 | 82.42 | 78.79 | 86.79 |
| CF-3D CNN (2022) | MRI+PET | 77.65 | 71.16 | 85.53 | 78.35 | 84.67 |
| Aging-Transformer (2023)* | MRI | 73.60 | 59.10 | 82.10 | 70.60 | 73.70 |
| DAUF (2023) | MRI | 75.32 | 70.65 | 82.63 | 76.64 | 81.95 |
| MMCA (2023) | MRI+PET+Structured | 78.12 | 72.87 | 86.07 | 79.47 | 86.05 |
| AFSNet (2024)* | Multiple MRI | 78.74 | **78.30** | 79.75 | 79.03 | 79.03 |
| SGCN (2025) | MRI+PET | 78.42 | 76.75 | 82.48 | 79.62 | 84.59 |
| MF-Transformer (2025) | MRI+PET+Structured | 79.69 | 75.22 | 88.14 | 81.68 | 86.90 |
| **Ours-1** | MRI+PET | 80.21 | 71.19 | 92.61 | 81.90 | 88.13 |
| **Ours-2** | MRI+PET+Structured | **80.71** | 72.73 | **92.68** | **82.71** | **89.70** |

BRC-MMHF reports two results: **Ours-1** uses MRI and PET only, whereas **Ours-2** extends this by incorporating the structured clinical data branch. For methods marked with \*, the source code and data were not released, and the metrics are directly taken from the original publication.

### D. Ablation Experiments

We conducted two ablation experiments to evaluate the effectiveness of different components of the BRC-MMHF framework. First, the impact of the channel exchange mechanism, hypergraph convolution, and hypergraph attention on feature fusion and classification performance is examined; Second, MRI, PET, and structured phenotypic data are removed individually to assess their contributions to MCI conversion diagnosis.

#### 1) Module Ablation

Four ablation models are designed to assess module-level effects: (1) **Model**: full model; (2) **Model 1**: no hypergraph attention (Hattn), disabling dynamic brainregion feature selection; (3) **Model 2**: no hypergraph convolution (Hgc), removing inter-region relationship modeling; (4) **Model 3**: no cross-modal channel exchange (CE), removing feature exchange between MRI and PET during encoding; (5) **Model 4**: no shared bottleneck layer (SL), decoding branches independently and concatenating features only at the classifier, reducing high-level semantic alignment.

As shown in Table V(a), removing Hattn and Hgc modules reduced ACC to 78.57% and 76.47% and AUC to 89.09% and 87.42%, indicating that brain-region-level hypergraph modeling makes an important contribution to the overall performance of the framework. Furthermore, removing the CE module lowered ACC to 76.32% and AUC to 87.0%, and removing the SL further reduced ACC to 75.42%. This indicates that CE helps align MRI and PET features and mitigate modality heterogeneity, while SL provides stable shared features.

**TABLE 5:** Ablation experiment results.

(a) Module ablation
| Model | SL | CE | Hgc | Hattn | ACC(%) ↑ | AUC(%) ↑ |
| --- | --- | --- | --- | --- | --- | --- |
| Model | ✓ | ✓ | ✓ | ✓ | **80.71** | **89.70** |
| Model 1 | ✓ | ✓ | ✓ |  | 78.57 | 89.09 |
| Model 2 | ✓ | ✓ |  |  | 76.47 | 87.42 |
| Model 3 | ✓ |  |  |  | 76.32 | 87.00 |
| Model 4 |  |  |  |  | 75.42 | 86.58 |

(b) Modality Ablation
| Model | MRI | PET | Table | ACC(%) ↑ | AUC(%) ↑ |
| --- | --- | --- | --- | --- | --- |
| Model M | ✓ |  |  | 67.43 | 68.51 |
| Model P |  | ✓ |  | 68.89 | 70.35 |
| Model T |  |  | ✓ | 78.12 | 87.16 |
| Model M+P | ✓ | ✓ |  | 80.21 | 88.13 |
| Model M+P+T | ✓ | ✓ | ✓ | **80.71** | **89.70** |

(a) Module Ablation

Overall, all submodules improved performance, confirming the modular design’s effectiveness.

#### 2) Modality Ablation

To further evaluate the contribution of each modality branch, modality ablation experiments are conducted: (1) **Model M**: using only MRI data, with deep features extracted by the encoder and fed into the classifier; (2) **Model P**: using only PET data, following the same process as (1); (3) **Model T**: using only tabular data; (4) **Model M+P**: removing the final-stage concatenation with tabular data and classifying using only the fused image features from the framework; (5) **Model M+P+T**: using all image and tabular data. The results are shown in Table V(b).

Using only MRI or PET resulted in low ACC and AUC, indicating limited effectiveness of single imaging modalities to simply extract whole-brain features for MCI classification. Tabular data alone performed better (ACC 78.12%) due to inclusion of clinically relevant scales such as FAQ and ADAS. Combining different modalities improved performance, with MRI-PET fusion (Model M+P) achieving 80.21% ACC and 88.13% AUC, confirming the advantage of the proposed BRC-MMHF framework.

In summary, ablation results show that each BRC-MMHF component is necessary: Channel exchange tightens MRI-PET semantic coupling, while hypergraph convolution and attention sharpen region-level modeling and capture higher-order relations; collaboratively, they form a full pipeline from interaction/alignment to structural modeling, boosting fusion performance. Modality ablations show that removing any imaging modality or clinical phenotype degrades results, whereas integrating imaging with clinical data improves robustness and interpretability. Overall, the findings validate the design and underscore its clinical potential.

### E. Interpretability Analysis

To assess the interpretability of the BRC-MMHF framework, we incorporated the Lesion-Aware Node Scoring (LANS) module during inference and used Eq. (14) to quantify brain region contributions. For each subject, brain regions are ranked by the *β*_*v*_, where the top 10 most influential regions are identified. Figure 5(a) shows these regions for two subjects, and Fig. 5(b) shows the 10 most discriminative regions between sMCI and pMCI identified by LANS. Using BrainNet Viewer [45], we visualized regions such as the left/right hippocampus, parahippocampal gyrus, left inferior temporal gyrus, and left inferior parietal lobule, which play critical roles in MCI conversion. These interpretability results help clinicians focus on high-risk regions, improve personalized follow-up and intervention strategies, and enhance the model’s clinical applicability.

**Fig. 5:**
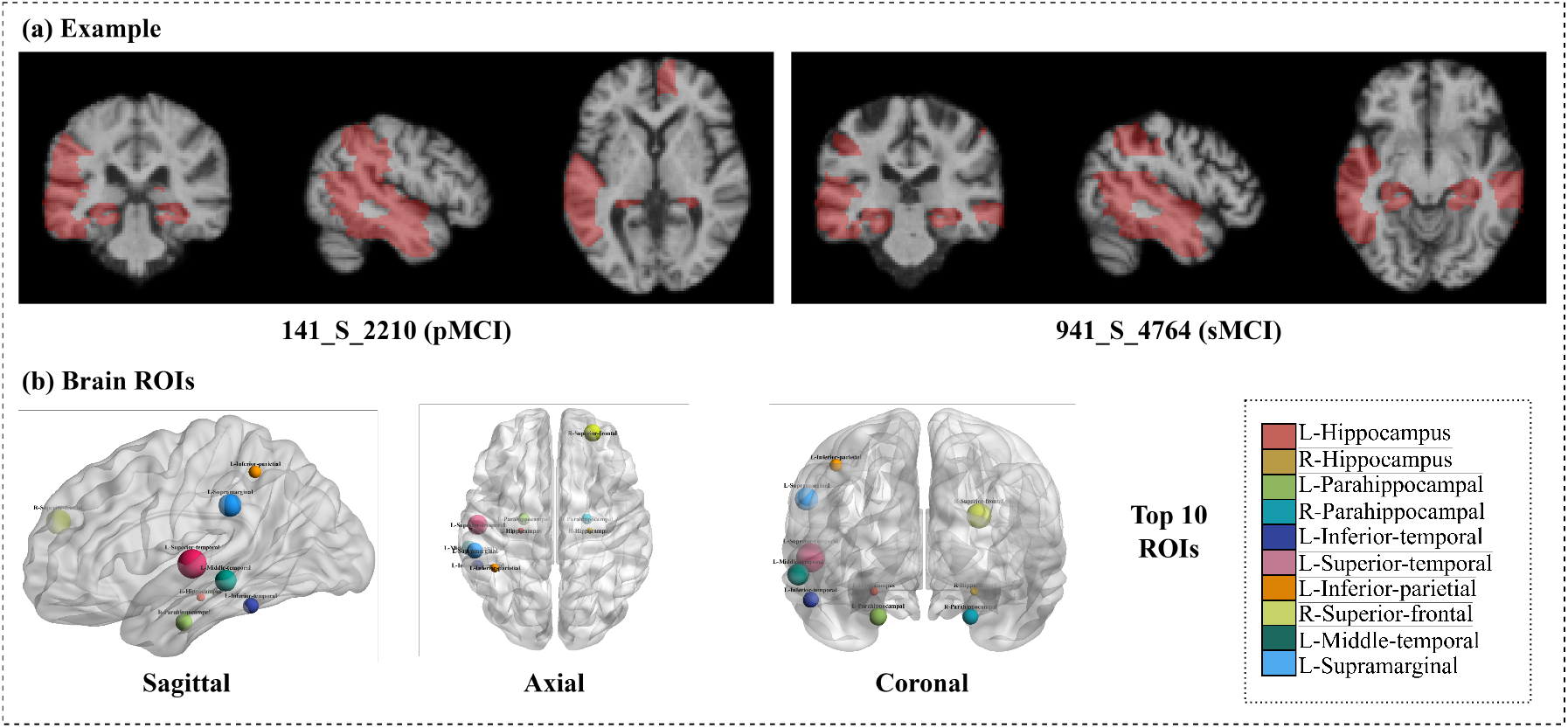
Interpretability analysis: (a) visualization of key brain regions in participants; (b) identification of the most important brain regions for MCI progression prediction.

In addition, SHAP [46] is applied to analyze the importance of tabular features. Specifically, a TabPFN classifier is fit on all available samples for attribution/visualization(no CV metrics are derived from this model). First-order Shapley values are estimated for the positive class (pMCI) using the full dataset as the background distribution. For each subject *i* and feature *j*, the explainer returns a SHAP value *ϕ*_*ij*_; global importance is reported as the mean absolute value mean_*i*_ |*ϕ*_*ij*_ |. Features are ranked by this global importance. The top 10 clinical variables are reported in Table VI, while Fig. 6 visualizes the per-subject attribution distributions in the test set. It can be observed that the key variables for predicting MCI progression are cognitive test variables such as RAVLT IMMEDIATE, ADAS and LDELTOTAL, which provide guidance for the preliminary screening of MCI conversion risk to Alzheimer’s disease in clinical practice. Moreover, even for the top 10 most important variables, their |*ϕ*_*ij*_ | values are relatively small, indicating that MCI conversion prediction relies more on the combination of multiple variables rather than a single feature.

**TABLE 6:** Top 10 most important clinical variables identified by SHAP analysis.

| Variable | Description |
| --- | --- |
| RAVLT_IMMEDIATE | RAVLT (Rey Auditory Verbal Learning Test) immediate recall |
| FAQ | Functional Activities Questionnaire, measuring daily functional abilities |
| ADAS13 | The 13-item Alzheimer's Disease Cognitive Scale total score |
| APOE4 | Apolipoprotein E (with allele types 0, 1, and 2 representing different genetic variations) |
| LDELTOTAL | Logical memory delayed recall total score |
| ADASQ4 | Alzheimer's Disease Assessment Scale - Question 4 |
| ECOGSPMEM | Everyday Cognition - Study Partner Memory score |
| ADAS11 | Alzheimer's Disease Assessment Scale, 11-item version total score |
| MPACCTRAILSB | Trail Making Test B, typically recorded in seconds, assessing executive function impairment |
| MPACCDIGIT | Digit test score from the modified Preclinical Alzheimer Cognitive Composite |

**Fig. 6:**
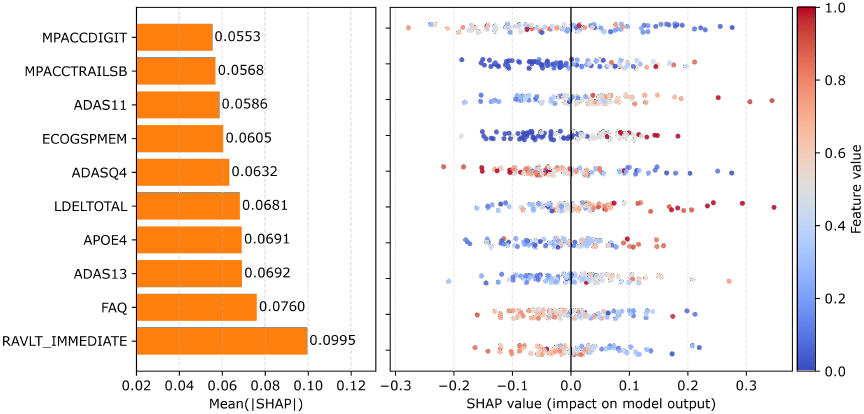
Interpretability analysis of structured clinical data (displaying only the top 10 variables).

### F. Summary of Findings

The experimental results highlight the strengths of the proposed BRC-MMHF framework across multiple dimensions. First, the framework demonstrated outstanding performance in terms of ACC, BACC, and AUC, outperforming recent approaches and achieving balanced effectiveness across the two classes, thereby ensuring reliable classification performance. Second, interpretability analysis confirmed that the LANS module effectively identified subject-specific lesion-related brain regions, including the hippocampus, parahippocampal gyrus, and inferior temporal gyrus. Moreover, SHAP analysis of the tabular branch showed that various clinical assessments and cognitive tests contributed differently to MCI conversion prediction, with the model’s decisions mainly driven by the joint influence of multiple measures rather than any single dominant factor. Third, ablation studies demonstrated that each module, channel exchange for modality alignment, hypergraph convolution and attention for high-order interregional modeling, and the tabular branch for phenotypic integration—contributed meaningfully, with their combination delivering the best performance. In conclusion, these results highlight BRC-MMHF’s ability to integrate predictive accuracy, interpretability, and modular flexibility, supporting its clinical applicability.

## V. Discussion

This study proposes the BRC-MMHF framework, which exhibits significant advantages in MCI conversion prediction and demonstrates strong capabilities in multimodal image fusion, while maintaining interpretability. From a clinical perspective, the proposed BRC-MMHF framework demonstrates strong practicality and tangible clinical relevance. First, the framework is consistent with the clinical diagnostic workflow for MCI and AD, and its ability to achieve accurate and efficient MCI conversion prediction within a 36-month window aligns well with current clinical practice. Application of BRC-MMHF integrated medical supporting software for MCI patient management and risk assessment enables clinicians to efficiently stratify patients into high and low risk groups, and initiate preventive or disease-modifying interventions in a timely manner. This is particularly important, as early treatment and lifestyle adjustments have been shown to delay cognitive decline and improve patients’ quality of life. Second, while maintaining high predictive accuracy, the framework also provides visual interpretability of its results. Compared with existing predictive methods, this capability offers evidence-based explanations that enhance readability and usability, thereby strengthening physician confidence and facilitating seamless integration into routine clinical workflows. Beyond prediction accuracy, the clinical impact of BRC-MMHF lies in its potential to improve patient management and healthcare resource allocation. The framework’s interpretability supports multidisciplinary collaboration among neurologists, radiologists, and caregivers, fostering consensus in treatment strategies. Moreover, by generating patient-specific risk assessments and highlighting lesion-relevant brain regions, the model can guide personalized follow-up schedules, enable more efficient use of clinical resources.

In addition, the framework also has potential to be applied for more generic brain disease prediction scenarios. Beyond MCI conversion prediction, BRC-MMHF can be extended to MCI and AD diagnosis as well as other brain diseases that involve MRI, PET, and structured clinical data.

Although the framework performs well in MCI conversion prediction, it has limitations. First, its generalization ability is constrained under small-sample conditions, as highquality annotated data—especially longitudinal data at the MCI stage—are scarce. Future work should explore transfer learning and related strategies to address this issue. Second, the imaging and non-imaging features lack deep integration, limiting the exploitation of their complementarity. Developing more advanced cross-modal interaction mechanisms may further enhance the characterization of disease mechanisms and improve predictive performance.

Future work will evaluate BRC-MMHF in longitudinal follow-up, multi-center transfer learning, and broader neurodegenerative disease settings. We will explore domain adaptation, cross-disease transfer, and temporal learning to improve robustness on multi-center, small-sample, and longitudinal cohorts. Although validated only for MCI conversion, its region-level modeling and high-order fusion paradigm may be generalizable to Parkinson’s disease and other dementias, such as frontotemporal dementia and vascular dementia.

## VI. Conclusion

This study proposes the BRC-MMHF framework to enhance multimodal fusion and improve interpretability in MCI conversion prediction. It integrates MRI and PET data, introduces parameter-free channel exchange in the shared encoder layer to reduce modality heterogeneity, and utilizes ROI-based feature extraction to obtain aligned brain region features. Hypergraph convolution and attention model high-order interregional relationships, while a lesion-aware module identifies key regions to enhance interpretability. A structured clinical data branch with a lightweight tabular encoder is integrated to improve adaptability and prediction accuracy. Experiments on the ADNI dataset show that BRC-MMHF outperforms several recent methods in sMCI vs. pMCI classification, demonstrating its potential for early AD diagnosis.

## Data Availability

All data produced are available online at

https://adni.loni.usc.edu/

## STATEMENTS

### A. Ethics Statement

This study utilized data from the Alzheimer’s Disease Neuroimaging Initiative (ADNI). All participants gave written informed consent, and neuroimaging and clinical data were provided by external centers in accordance with ethical standards and de-identified before release. As only publicly available data were reanalyzed, no further ethical approval was required.

### B. Data Availability Statement

Data are available from the ADNI website (https://adni.loni.usc.edu/) and the USC IDA ARC Builder (IDs: ADNI1, ADNIGO, ADNI2), following standard multimodal AD protocols for comparability and reproducibility.

## References

[1] Z. Breijyeh and R. Karaman, “Comprehensive review on alzheimer’s disease: causes and treatment,” Molecules, vol. 25, no. 24, p. 5789, 2020.

[2] R. Brookmeyer, E. Johnson, K. Ziegler-Graham, and H. M. Arrighi, “Forecasting the global burden of alzheimer’s disease,” Alzheimer’s & dementia, vol. 3, no. 3, pp. 186–191, 2007.

[3] G. M. McKhann, D. S. Knopman, H. Chertkow, B. T. Hyman, C. R. Jack Jr, C. H. Kawas, W. E. Klunk, W. J. Koroshetz, J. J. Manly, R. Mayeux et al., “The diagnosis of dementia due to alzheimer’s disease: recommendations from the national institute on aging-alzheimer’s association workgroups on diagnostic guidelines for alzheimer’s disease,” Alzheimer’s & dementia, vol. 7, no. 3, pp. 263–269, 2011.

[4] A. J. Mitchell and M. Shiri-Feshki, “Rate of progression of mild cognitive impairment to dementia–meta-analysis of 41 robust inception cohort studies,” Acta psychiatrica scandinavica, vol. 119, no. 4, pp. 252–265, 2009.

[5] J. Whitwell, K. Josephs, M. Murray, K. Kantarci, S. Przybelski, S. Weigand, P. Vemuri, M. Senjem, J. Parisi, D. Knopman et al., “Mri correlates of neurofibrillary tangle pathology at autopsy: a voxel-based morphometry study,” Neurology, vol. 71, no. 10, pp. 743–749, 2008.

[6] A. Rogeau, F. Hives, C. Bordier, H. Lahousse, V. Roca, T. Lebouvier, F. Pasquier, D. Huglo, F. Semah, and R. Lopes, “A 3d convolutional neural network to classify subjects as alzheimer’s disease, frontotemporal dementia or healthy controls using brain 18f-fdg pet,” Neuroimage, vol. 288, p. 120530, 2024.

[7] S. Basaia, F. Agosta, L. Wagner, E. Canu, G. Magnani, R. Santangelo, M. Filippi, A. D. N. Initiative et al., “Automated classification of alzheimer’s disease and mild cognitive impairment using a single mri and deep neural networks,” NeuroImage: Clinical, vol. 21, p. 101645, 2019.

[8] J. Zhang, B. Zheng, A. Gao, X. Feng, D. Liang, and X. Long, “A 3d densely connected convolution neural network with connection-wise attention mechanism for alzheimer’s disease classification,” Magnetic Resonance Imaging, vol. 78, pp. 119–126, 2021.

[9] Z. Zhang, L. Gao, P. Li, G. Jin, J. Wang, A. D. N. Initiative et al., “Dauf: A disease-related attentional unet framework for progressive and stable mild cognitive impairment identification,” Computers in Biology and Medicine, vol. 165, p. 107401, 2023.

[10] J. Young, M. Modat, M. J. Cardoso, A. Mendelson, D. Cash, S. Ourselin, A. D. N. Initiative et al., “Accurate multimodal probabilistic prediction of conversion to alzheimer’s disease in patients with mild cognitive impairment,” NeuroImage: Clinical, vol. 2, pp. 735–745, 2013.

[11] Y. Huang, J. Xu, Y. Zhou, T. Tong, X. Zhuang, and A. D. N. I. (ADNI), “Diagnosis of alzheimer’s disease via multi-modality 3d convolutional neural network,” Frontiers in neuroscience, vol. 13, p. 509, 2019.

[12] M. Odusami, R. Maskeliūnas, R. Damaševičius, and S. Misra, “Explainable deep-learning-based diagnosis of alzheimer’s disease using multimodal input fusion of pet and mri images,” Journal of Medical and Biological Engineering, vol. 43, no. 3, pp. 291–302, 2023.

[13] Z. Kong, M. Zhang, W. Zhu, Y. Yi, T. Wang, and B. Zhang, “Multimodal data alzheimer’s disease detection based on 3d convolution,” Biomedical Signal Processing and Control, vol. 75, p. 103565, 2022.

[14] J. Zhang, X. He, Y. Liu, Q. Cai, H. Chen, and L. Qing, “Multi-modal cross-attention network for alzheimer’s disease diagnosis with multimodality data,” Computers in biology and medicine, vol. 162, p. 107050, 2023.

[15] Y. Zhang, K. Sun, Y. Liu, F. Xie, Q. Guo, and D. Shen, “A modality-flexible framework for alzheimer’s disease diagnosis following clinical routine,” IEEE Journal of Biomedical and Health Informatics, 2024.

[16] Y. Zhu, J. Ma, C. Yuan, and X. Zhu, “Interpretable learning based dynamic graph convolutional networks for alzheimer’s disease analysis,” Information Fusion, vol. 77, pp. 53–61, 2022.

[17] B. Lei, Y. Li, W. Fu, P. Yang, S. Chen, T. Wang, X. Xiao, T. Niu, Y. Fu, S. Wang et al., “Alzheimer’s disease diagnosis from multi-modal data via feature inductive learning and dual multilevel graph neural network,” Medical Image Analysis, vol. 97, p. 103213, 2024.

[18] H. Zhou, L. He, B. Y. Chen, L. Shen, and Y. Zhang, “Multi-modal diagnosis of alzheimer’s disease using interpretable graph convolutional networks,” IEEE Transactions on Medical Imaging, 2024.

[19] I. Arevalo-Rodriguez, N. Smailagic, M. R. i Figuls, A. Ciapponi, E. Sanchez-Perez, A. Giannakou, O. L. Pedraza, X. B. Cosp, and S. Cullum, “Mini-mental state examination (mmse) for the detection of alzheimer’s disease and other dementias in people with mild cognitive impairment (mci),” Cochrane database of systematic reviews, no. 3, 2015.

[20] R. Cuingnet, E. Gerardin, J. Tessieras, G. Auzias, S. Lehéricy, M.-O. Habert, M. Chupin, H. Benali, O. Colliot, A. D. N. Initiative et al., “Automatic classification of patients with alzheimer’s disease from structural mri: a comparison of ten methods using the adni database,” neuroimage, vol. 56, no. 2, pp. 766–781, 2011.

[21] C.-Y. Wee, P.-T. Yap, D. Shen, and A. D. N. Initiative, “Prediction of alzheimer’s disease and mild cognitive impairment using cortical morphological patterns,” Human brain mapping, vol. 34, no. 12, pp. 3411–3425, 2013.

[22] Z. Sun, Y. Qiao, B. P. Lelieveldt, M. Staring, A. D. N. Initiative et al., “Integrating spatial-anatomical regularization and structure sparsity into svm: Improving interpretation of alzheimer’s disease classification,” NeuroImage, vol. 178, pp. 445–460, 2018.

[23] I. Beheshti, H. Demirel, H. Matsuda, A. D. N. Initiative et al., “Classification of alzheimer’s disease and prediction of mild cognitive impairment-to-alzheimer’s conversion from structural magnetic resource imaging using feature ranking and a genetic algorithm,” Computers in biology and medicine, vol. 83, pp. 109–119, 2017.

[24] E. Westman, J.-S. Muehlboeck, and A. Simmons, “Combining mri and csf measures for classification of alzheimer’s disease and prediction of mild cognitive impairment conversion,” Neuroimage, vol. 62, no. 1, pp. 229–238, 2012.

[25] N. Jia, T. Jia, L. Zhao, B. Ma, and Z. Zhu, “Multi-modal global-and local-feature interaction with attention-based mechanism for diagnosis of alzheimer’s disease,” Biomedical Signal Processing and Control, vol. 95, p. 106404, 2024.

[26] M. Luo, Z. He, H. Cui, Y.-P. P. Chen, P. Ward, A. D. N. Initiative et al., “Class activation attention transfer neural networks for mci conversion prediction,” Computers in Biology and Medicine, vol. 156, p. 106700, 2023.

[27] L. Liu, S. Liu, L. Zhang, X. V. To, F. Nasrallah, and S. S. Chandra, “Cascaded multi-modal mixing transformers for alzheimer’s disease classification with incomplete data,” NeuroImage, vol. 277, p. 120267, 2023.

[28] Q. Yu, Q. Ma, L. Da, J. Li, M. Wang, A. Xu, Z. Li, W. Li, A. D. N. Initiative et al., “A transformer-based unified multimodal framework for alzheimer’s disease assessment,” Computers in Biology and Medicine, vol. 180, p. 108979, 2024.

[29] Z. Chen, Y. Liu, Y. Zhang, Q. Li, A. D. N. Initiative et al., “Orthogonal latent space learning with feature weighting and graph learning for multimodal alzheimer’s disease diagnosis,” Medical Image Analysis, vol. 84, p. 102698, 2023.

[30] Y. Jiang, Y. Gao, Z. Zhu, C. Yan, and Y. Gao, “Hyperrep: Hypergraph-based self-supervised multimodal representation learning,” 2024.

[31] Z. Qiu, P. Yang, C. Xiao, S. Wang, X. Xiao, J. Qin, C.-M. Liu, T. Wang, and B. Lei, “3d multimodal fusion network with disease-induced joint learning for early alzheimer’s disease diagnosis,” IEEE Transactions on Medical Imaging, vol. 43, no. 9, pp. 3161–3175, 2024.

[32] R. R. Selvaraju, M. Cogswell, A. Das, R. Vedantam, D. Parikh, and D. Batra, “Grad-cam: Visual explanations from deep networks via gradient-based localization,” in Proceedings of the IEEE international conference on computer vision, 2017, pp. 618–626.

[33] J. Liu, Y. Xu, Y. Liu, H. Luo, W. Huang, and L. Yao, “Attention-guided 3d cnn with lesion feature selection for early alzheimer’s disease prediction using longitudinal smri,” IEEE Journal of Biomedical and Health Informatics, 2024.

[34] D. Gunning, M. Stefik, J. Choi, T. Miller, S. Stumpf, and G.-Z. Yang, “Xai—explainable artificial intelligence,” Science robotics, vol. 4, no. 37, p. eaay7120, 2019.

[35] Ö. Çiçek, A. Abdulkadir, S. S. Lienkamp, T. Brox, and O. Ronneberger, “3d u-net: learning dense volumetric segmentation from sparse annotation,” in International conference on medical image computing and computer-assisted intervention. Springer, 2016, pp. 424–432.

[36] P.-N. Bui, D.-T. Le, J. Bum, and H. Choo, “Multi-scale feature enhancement in multi-task learning for medical image analysis,” arXiv preprint 2412.00351, 2024.

[37] N. Tishby and N. Zaslavsky, “Deep learning and the information bottleneck principle,” in 2015 ieee information theory workshop (itw). Ieee, 2015, pp. 1–5.

[38] Z. Marinov, S. Reiß, D. Kersting, J. Kleesiek, and R. Stiefelhagen, “Mirror u-net: Marrying multimodal fission with multi-task learning for semantic segmentation in medical imaging,” in Proceedings of the IEEE/CVF International Conference on Computer Vision, 2023, pp. 2283–2293.

[39] N. Tzourio-Mazoyer, B. Landeau, D. Papathanassiou, F. Crivello, O. Etard, N. Delcroix, B. Mazoyer, and M. Joliot, “Automated anatomical labeling of activations in spm using a macroscopic anatomical parcellation of the mni mri single-subject brain,” Neuroimage, vol. 15, no. 1, pp. 273–289, 2002.

[40] Q. Zuo, B. Lei, Y. Shen, Y. Liu, Z. Feng, and S. Wang, “Multi-modal representations learning and adversarial hypergraph fusion for early alzheimer’s disease prediction,” in Chinese Conference on Pattern Recognition and Computer Vision (PRCV). Springer, 2021, pp. 479– 490.

[41] Y. Gao, Y. Feng, S. Ji, and R. Ji, “Hgnn+: General hypergraph neural networks,” IEEE Transactions on Pattern Analysis and Machine Intelligence, vol. 45, no. 3, pp. 3181–3199, 2022.

[42] S. Bai, F. Zhang, and P. H. Torr, “Hypergraph convolution and hypergraph attention,” Pattern Recognition, vol. 110, p. 107637, 2021.

[43] N. Hollmann, S. Müller, L. Purucker, A. Krishnakumar, M. Körfer, S. B. Hoo, R. T. Schirrmeister, and F. Hutter, “Accurate predictions on small data with a tabular foundation model,” Nature, vol. 637, no. 8045, pp. 319–326, 2025.

[44] X. Gao, H. Cai, and M. Liu, “A hybrid multi-scale attention convolution and aging transformer network for alzheimer’s disease diagnosis,” IEEE Journal of Biomedical and Health Informatics, vol. 27, no. 7, pp. 3292– 3301, 2023.

[45] M. Xia, J. Wang, and Y. He, “Brainnet viewer: a network visualization tool for human brain connectomics,” PloS one, vol. 8, no. 7, p. e68910, 2013.

[46] S. M. Lundberg and S.-I. Lee, “A unified approach to interpreting model predictions,” in Advances in Neural Information Processing Systems (NeurIPS), vol. 30, 2017.

